# Assessment of a combined manual therapy and taping method for the treatment of chronic lower back pain A randomized controlled trial

**DOI:** 10.1101/2020.03.19.20024950

**Authors:** Stefan Schmidt, Nicolas Keim, Claudia Schultz, Dieter Sielmann, Roman Huber, Harald Walach

**Author notes:** **Corresponding Author** Prof. Dr. phil. Stefan Schmidt, Dept. for Psychosomatic Medicine and Psychotherapy, Medical Faculty, Medical Center – University of Freiburg, Hauptstr. 8, 79104 Freiburg, Germany.

## Abstract

**Background:** Chronic lower back pain is the most frequent medical problem and the condition with the most years lived with disability. A pragmatic RCT was performed to assess a new treatment, Medi-Taping, which aims at reducing complaints by treating pelvic obliquity with a combination of manual treatment of trigger points and kinesio taping.

**Methods:** 110 patients were randomized at two study centers either to Medi-Taping or to a standard treatment consisting of psychoeducation and physiotherapy as control. Treatment duration was three weeks. Measures were taken at baseline, end of treatment and at follow-up after two months. Main outcome criteria were lower back pain measured with VAS, the Chronic Pain Grade Scale and the Oswestry Low Back Pain Disability Questionnaire.

**Results:** Patients of both groups benefited from the treatment by medium to large effect sizes. All effects were pointing towards the intended direction with patients receiving Medi-Taping doing better. But at end of treatment and follow-up there were no significant differences for the primary endpoints between groups. Health related quality of life was significantly higher (p=.004) in patients receiving Medi-Taping compared to controls.

**Conclusions:** Medi-Taping, a purported way of correcting pelvic obliquity and chronic tension resulting from it, is a treatment modality similar in effectiveness as a complex physiotherapy and patient education program.

**Significance:** This RCT evaluated the effect of a combined therapy consisting of manual treatment and kinesio tape in patients with lower back pain. Patients receiving this treatment benefitted substantially but so did patients in the active control condition receiving physiotherapy and patient education. However, patients receiving the combined treatment had a significant higher quality of life.

## Introduction

### Background

Chronic back pain (CLBP) is one of the most frequent medical problems in Western countries. Lifetime prevalence rates of about 75% and point prevalence rates between 32% and 49% have been documented by epidemiological studies in Germany (Raspe 2012; Schmidt et al., 2007) and are similar elsewhere in Europe (Ekman et al., 2005; Farioli et al., 2014; Spijker-Huiges et al., 2015). Worldwide, it is the condition with the most years lived with disability (Global Burden of Disease Study 2013 Collaborators et al., 2015). Although we have powerful medications to treat acute pain conditions, for chronic conditions most guidelines are meanwhile advising against medications, because of side-effects in long-term users even of simple nonsteroidal anti-inflammatory substances, because of a lack of clinical effectiveness and because of potential dependency problems with stronger opiates (Coxib and traditional NSAID Trialists’ (CNT) Collaboration 2013; Dowell et al., 2016; Gor and Saksena 2011; Kuijpers et al., 2011). Thus, chronic low-back pain still poses a therapeutic challenge to general practitioners who are the first in line for patients afflicted with CLBP. Physiotherapy, including mobilization, as well as exercise, Yoga, Tai Chi and other forms of complex modalities all have shown some efficacy for CLBP (Chou et al., 2017; Qaseem et al., 2017; Saper et al., 2017; Tilbrook et al., 2011; Wellington 2014), yet some patients either remain unimproved even after complex applications or standard therapies, or are unwilling to undergo such complex and comparatively demanding therapeutic regimes as Yoga therapy or exercise programs.

The aetiology of CLBP is unclear in most cases, although we know in general terms that in some patients central hypersensitization plays a role as well as pain memory on the level of the spinal cord ganglia. But in most cases many factors contribute to the generation and to sustaining a chronic pain problem (Nijs et al., 2017). Local or distal tensions are likely contributors, and ultrasound data point to the fact that connective tissue alterations might be the cause for pain (Langevin et al., 2009). Such connective tissue alterations are likely the long term-sequelae of past traumata (Ofner et al., 2017), chronic postural or functional muscular problems (Bordoni and Marelli 2017; Ingber et al., 2014; Schleip et al., 2012; Tozzi 2015a; b). A neglected kind of potential causal problems are postural ones such as pelvic obliquity. In this under-researched area very few studies have documented this potential problem (Ayanniyi et al., 2008).

This was one of the starting points for our therapeutic rationale: In the clinical experience of one of the authors (DS) pelvic obliquity might be a potential co-factor in a multicausal network of factors contributing to the genesis of CLBP, resulting in chronic tension and painful trigger points that radiate out or induce pain through secondary reflex systems such as the Head zones (Tozzi 2015a; b). Thus, he developed a method of treating CLBP using a certain kind of kinesio tape, but in a modified pattern. While classical kinesio tape, developed by Kenzo Kase in the 60ies, mainly to treat and prevent sports injuries (Kase et al., 1996), has become a standard treatment in sports medicine, despite mixed and unclear effects of its clinical effectiveness (Ilic et al., 2017; Lim and Tay 2015; Nelson 2016; Parreira et al., 2014; Vanti et al., 2015; Williams et al., 2012), the way it was used in our study is contingent on DS’s model of pelvic obliquity and secondary trigger points. Thus, in our treatment model those trigger points are treated by acupressure and massage first and the muscular reflex patterns disrupted by placing kinesio tape along those muscles that are thought to be responsible for triggering and supporting the pain processes. This combinded treatment approach is termed *Medi-Taping*.

Since this particular type of kinesio taping as a supportive treatment for correcting pelvic obliquity as a potential cofactor in causing CLBP has never been studied, we designed a pragmatic pilot evaluation study to see whether this modality would warrant further evaluation.

### Objectives

The objective of this pragmatic, randomized controlled trial thus was to study the clinical effects of correcting pelvic obliquity and trigger points with short acupressure and massage and applying kinesio tape to the respective muscles. We did this by comparing this new modality to best-practice physiotherapy, a strong kind of control which is guideline supported, as there is sufficient evidence for its effectiveness in CLBP (Critchley et al., 2007; van Middelkoop et al., 2011). Since we did not know what effect sizes were to be expected for the medi-taping condition and in what domains potential effects would show we opted for a broad array of pain and functional measurements following respective recommendations (Dworkin et al., 2005). Since this was a first study, we defined several primary and secondary outcomes. Because DS has seen strong improvements using this method in his GP clinic, we opted for a superiority design.

## Materials and Methods

### Design

We conducted a pragmatic, assessor blinded, randomized controlled trial at two different study centers. Patients with CLBP were randomly allocated to either Medi-Taping or a standard treatment for CLPB consisting of psychoeducation and physiotherapy for three weeks. Measurements were taken at baseline (t^1^), after the end of the respective treatment (t^2^, 4 weeks after baseline) and at follow-up two months after the end of treatment (t^3,^ three months after baseline).

### Patients

#### Inclusion Criteria

Criteria for the inclusion in our trial were unspecific lower back pain for more than 12 weeks, a rating of at least 4 cm on a 10 cm Visual Analog Scale (VAS) for the back pain, age between 18 and 80 years, and the ability to read and communicate in German.

#### Exclusion Criteria

Patients fulfilling one of the following criteria were excluded from the trial: neurological malfunction at the lower extremities related to CLBP, a rating larger than 8cm on the VAS for back pain, back pain due to infection, tumor, osteoporosis, stenosis of the cerebrospinal canal, slipped vertebra, vertebral fractures, spinal disc herniation, surgery of vertebra, spinal disc or sacroiliac joint. Furthermore allergy to tape, pregnancy, current cancer diagnosis, addictive disorder, severe psychiatric disorder, significant impairment due to memory problems or brain disorders, artificial hip ankle or knee joint, participations in other RCTs.

#### Recruitment

Patients were recruited by public information, i.e. a press release of the Medical Center of the University of Freiburg, newspaper articles, information on the intranet of the Medical Center, information leaflets in pharmacies as well as two radio interviews at both study centers. Additionally, the GP clinic of DS in the study center Bad Oldesloe is well known for lower back pain treatment and attracted many patients by itself.

#### Ethics

The RCT was approved by the ethics committee of the Medical Center, University of Freiburg. All patients gave written informed consent before inclusion into the trial.

#### Study settings

The study was conducted at two different settings in Germany. One setting was the Outpatient Center for Complementary Medicine at the Medical Center of the University of Freiburg in the south of Germany (Study Center South). Patients allocated to Medi-Taping were treated within the outpatient center while Patients randomized to physiotherapy were treated in a larger private physiotherapy center (Reha Süd GmbH, Freiburg, Germany). The second setting was the clinic for general medicine of the author DS in Bad Odesloe in the northern part of Germany (Study Center North). Here patients allocated to the MediTaping were treated within the clinic of DS, patients receiving physiotherapy were sent to a physiotherapy center (Reha Aktiv, Bad Oldesloe, Germany)

#### Intervention

All patients received an active treatment for three weeks. This was either Medi-Taping or a standard treatment combined of psychoeducation and physiotherapy.

#### Medi-Taping

Patients received three treatments within three weeks on average, i.e. one treatment per week. The procedure within a session was standardized and manualized. According to this protocol the sessions started with an assessment of leg length difference. Patients were asked to lie on the back and the legs were slightly stretched by a soft pull at the ankles. Next a continuous horizontal line was drawn on the inside of both calves indicating the position of the calves towards each other. Then the patient was asked to sit up with the legs remaining stretched. This procedure results in a shift of the line between the two calves for most people. This shift was measured in millimeters as leg length difference.

The patient was then asked to stretch out, lying supine, and the therapist palpated any myogeloses and tense muscles areas that could be found next to the cervical spine between skullcap and seventh cervical vertebra from both sides. After this treatment the leg length difference assessment was repeated. If there was still a substantial difference the same treatment was also performed on the thoracic and lumbar spine. Also the mandibular joint was assessed for tense muscles and if necessary treated by palpation.

Next, the leg length difference was assessed again and several tapes were applied as follows. At first two parallel tapes were fixed on both sides of the spine above the erector spinae muscles ranging from the skullcap to the sacrum. For the application patients were asked to bend forward and to lean on the bench. This position stretches the back and its anatomical structure before applying the tape and thus provides the tape with tension before fixing it. Next a star-shaped tape (three stripes meeting in on point) was placed at the lower back while the patient was still in the same bending position. The star tape covered the area of the patient’s maximum pain and additionally stabilized the sacroiliac joint. This tape was placed with maximum tension in the middle part of the tape by stretching it before application while the ends of the tape (approx. 5 cm) were applied without tension. If after this procedure there was still residual pain, a third tape was placed at the gluteus maximus muscle. This tape was first fixed distally from the greater trochanter then stretched up to approx. 80 % of the possible tension before the other end was placed on the sacrum. On average six tapes were applied for the gluteus tape.

Tapes had to be kept as long as they stuck to the skin. If the patients had recurring LBP within the same week they were asked to see the therapist again immediately. Otherwise the second and the third treatment were scheduled for the following week respectively.

#### Standard Treatment

Patients allocated to the control or standard group received a booklet containing psychoeducative information for patients with lower back pain. This booklet was published by the commission for the German national guideline for the treatment of lower back pain (Ärztliches Zentrum für Qualität in der Medizin (ÄZQ) 2011). Furthermore they received a standard physiotherapy for CLBP, 6 sessions of 20 minute duration within three weeks. Physiotherapy was conducted by professional and commercial physiotherapy centers. They were paid for the treatment by the study center and provided standard documentation of the sessions conducted, but were not otherwise associated with the study.

#### Outcomes

The following outcome measures were applied.

VAS

LPB within the last two weeks was assessed by a Visual Analog Scale (VAS) of 10 cm length with the anchor points at 0cm ‘no pain at all’ and at 10cm ‘worst possible pain’ (Huskisson 1974; 1983).

#### Oswestry Low Back Pain Disability Questionnaire

Functional limitations due to CLBP was assesses by the Oswestry Low Back Pain Disability Questionnaire (ODQ) in its appropriate German version (Fairbank et al., 1980; Gaul et al., 2008). The ODQ is a self-completed questionnaire with ten items covering pain intensity, ability to care for oneself, lifting and carrying, ability to walk, ability to sit, ability to stand, sleep quality, social life, sexuality and ability to travel. Every item has six statements describing possible situations in the patient’s life. The most applicable statement is checked by the patient. Questions are scored by scale of 0-5.

We adapted the questionnaire by omitting one item regarding sexual function.

#### Chronic Pain Grade Scale/Korff Grading (CPGS)

This instrument consists of 6 numeric rating scales and a single question (Von Korff et al., 1992). The first item asks about activities of daily living and on how many days these could not be performed. Items two to four ask about current pain, worst pain and median pain during the last three months in a numeric format. Items five and six ask about the impact of pain on daily life and family/leisure activities over the last three months. These items are transformed into a grading of pain severity from one to four, with grade 1 and 2 reflecting low and grade 3 and 4 high disability. Cronbach’s alpha was reported to be .74 and thus acceptable in the original version and .88 in the German language version (Klasen et al., 2004).

#### Range of Motion: Fingertip to floor (FTF)

The distance between fingertips and floor (FTF) is a measure for the flexibility of the spine. It is measured by bending forward with stretched legs as far as possible. The distance between fingertips and floor is measured by a scale fixed to the wall. Individual scores have a limited validity but repeated measures are an appropriate clinical indicator for the mobility of the spine.

#### Schober Sign (SS)

The Schober Sign (SS) is another method to assess spinal mobility (Moll and Wright 1971), originally developed by Schober (Schober 1937). In order to assess the Schober Sign two marks are made on the skin overlying the lumbo-sacral spine while the patient stands erect. The first mark is made at the first processus of the sacrum, the second mark 10 cm cranially of the first mark. Next the patient is asked to bend as far as possible forward while the legs remain stretched. The distance between the two marks is measured in centimeters.

#### Leg length difference (LLD)

The procedure of measuring leg length difference is described above (see Treatment). A line indicating the position of two legs is drawn connecting both calves of a patient lying on the back. Next the patient is asked to sit up with the legs stretched. The shift between the two lines in this sitting position indicates the leg length difference which is an indicator of pelvic obliquity.

#### Quality of Life Profile for the Cronically Ill (PLC)

The Quality of Life Profile for the Chronically Ill is a health related quality of life inventory especially designed for patients with chronic conditions and validated in German (Siegrist et al., 1994). It consists of 40 items and 6 subscales: physical functioning, ability to relax and enjoy life, positive affect, negative affect, social contact, and social integration. Scores of the 6 subscales can be summed to a total score. The inventory is well validated and frequently used in the German speaking countries.

#### Primary and secondary outcomes

We predefined changes at t2 for pain (VAS and Korff pain grade), and functional limitations (Oswestry Disability Score) as primary outcomes. Secondary outcomes were changes in quality of life (PLC), spinal mobility (SS and DFF) and leg length difference as indicator of pelvic obliquity) at t2 and all changes at t3.

### Procedures

Patients contacted the study center and were screened for eligibility. Next they were invited to the respective study center and a clinical examination regarding inclusion/exclusion criteria was performed by CS (Freiburg) or DS (Bad Oldesloe). Patients were informed about the study and gave written informed consent. Having consented, patients filled in the questionnaires. After baseline measurements were completed patients received their group assignment. Patients assigned to Medi-Taping received their first treatment immediately after the assignment and were then scheduled for the next two weeks. Medi-Taping treatments lasted approx. 15 minutes. Patients assigned to standard treatment received the booklet on lower back pain and were connected with the respective physiotherapy center in order to schedule 6 sessions of physiotherapy of 20 min duration. Patients receiving pharmacotherapy for their back pain were allowed to continue with this treatment as they wished.

Patients returned to the study center one week after the end of treatment (t2) and were assessed again clinically (LLD, SS, DFF) and filled in questionnaires (ODQ, PLC, VAS, Korff). The same assessment was again at t3, approx. three months after baseline. Clinical examinations at t2 and t3 were made by trained examiners who had not been interacting with the patients and who were blinded against treatment allocation.

After follow-up measurements patients in the standard treatment arm were offered to receive Medi-Taping therapy if they so wished, free of charge.

### Power Analysis

There is no prior study for the estimation of an effect size of an approach combining manual therapy in order to reduce pelvic obliquity in combination with taping for CLBP. Thus, a clinical-pragmatic approach was taken. Based on reports of practitioners and patients a superiority effect of more than half a standard deviation was assumed. With an effect size of d= 0.6-0.7 and an objective to recruit at least 100 patients in total, a power range from 70% (d=0.6, α=.016, two-tailed, conservative correction for multiple testing) to 85% (d=0.7, α=.016, two-tailed) was achieved.

### Randomization and Allocation

Randomization was performed by using the random number generator of IBM SPSS 22 and allocation was blinded. Patients were randomized in blocks of 20. At first 5 Blocks of 20 were randomized at once. With ongoing recruitment a sixth block of 20 patients was randomized. Finally for the inclusion of the last patients a seventh block was randomized with only ten patients. Blocks were separated by study center. Study center North received 60 envelopes, South 70.The result of the randomization was printed on a result sheet that was sealed in an opaque envelope. The envelopes had consecutive numbers starting with one. The result sheet contained again the consecutive number, the group assignment and space to fill in date and time of opening, patient ID and name, name of person handing over the group assignment and their respective signature. Randomization was performed by SS who had otherwise no direct contact with patients. He ran the random number generator and sealed the envelopes that were then handed over to the respective clinician. The envelopes were opened in the presence of the respective patient in subsequent order. This process was documented by filling in the above result sheet.

### Blinding

All measurements at baseline took place before group assignment and were thus blinded. The patients themselves knew whether they received Medi-Taping or standard care and were thus not blinded. Measurements at t2 and t3 were performed by MDs who had no prior contact with the patients and who were blind to the group assignment. Patients were asked not to reveal any information regarding the therapy received towards the examiner in order to maintain the blinding.

### Statistical Methods

The study was evaluated according to the intention to treat approach. Missing data on questionnaire scales up to 20% of the total item number of the scale were replaced by means of the other items of the respective scale. All other missing data were replaced by regression-based imputations. Predictors for the imputation were the respective baseline value as well as age, gender, education, VAS at baseline, and chronic pain severity grade at t1.

For the baseline comparison of the two groups and the two study centers either t-test for independent data or Chi^2^-tests were applied. For the assessment of the primary and secondary outcome at t2 general linear models were applied with group and study center as dichotomous and baseline measure (t1) as continuous predictors. Data that followed a Poisson distribution (finger floor distance, Schober sign, Korff grading, difference in leg length) were evaluated with a linear model using a Poisson distribution, with baseline measure as continuous predictor and group and center as categorical predictors. Data that were continuous but deviated from a normal distribution (Oswestry Disability Questionnaire) were log-transformed prior to analysis. Since three primary outcome variables were applied we applied a correction for multiple testing according to Holm, which uses alpha/3 for the first criterion, alpha/2 for the second, alpha/1 for the third criterion (Holm 1979).

For the assessment of the follow-up data a repeated measurement ANOVA with t1, t3, and t3 as within-subject factor and *group assignment* and *study center* as between-subject factor was performed. Here the interaction term time × group was the focus of the analysis. Degrees of freedom were corrected for sphericity according to Greenhouse-Geisser. Regarding effect sizes we report *partial η^2^* from the respective analyses. However in order to compare our findings we also computed *Cohen’s d* for the changes from *baseline to end of therapy* (t1-t2) and from *baseline to follow-up* (t1-t3), by subtracting the means and dividing them to the mean of the respective standard deviations. All analyses were conducted with IBM SPSS 23 or Statistica V. 8.

## Results

### Recruitment and patient sample

Recruitment took place between June 2015 and March 2016 and was terminated when the specified number of patients was reached. Overall 561 patients were screened either per telephone or per email between June and October 2015. 147 patients were invited for a clinical examination, 119 patients showed up and finally 110 patients started with the intervention and were included into the intention to treat analysis. Of these 110 patients 59 were treated at the study center south and 51 at the study center north. The exact patient flow can be seen in figure 1.

**Figure 1.**
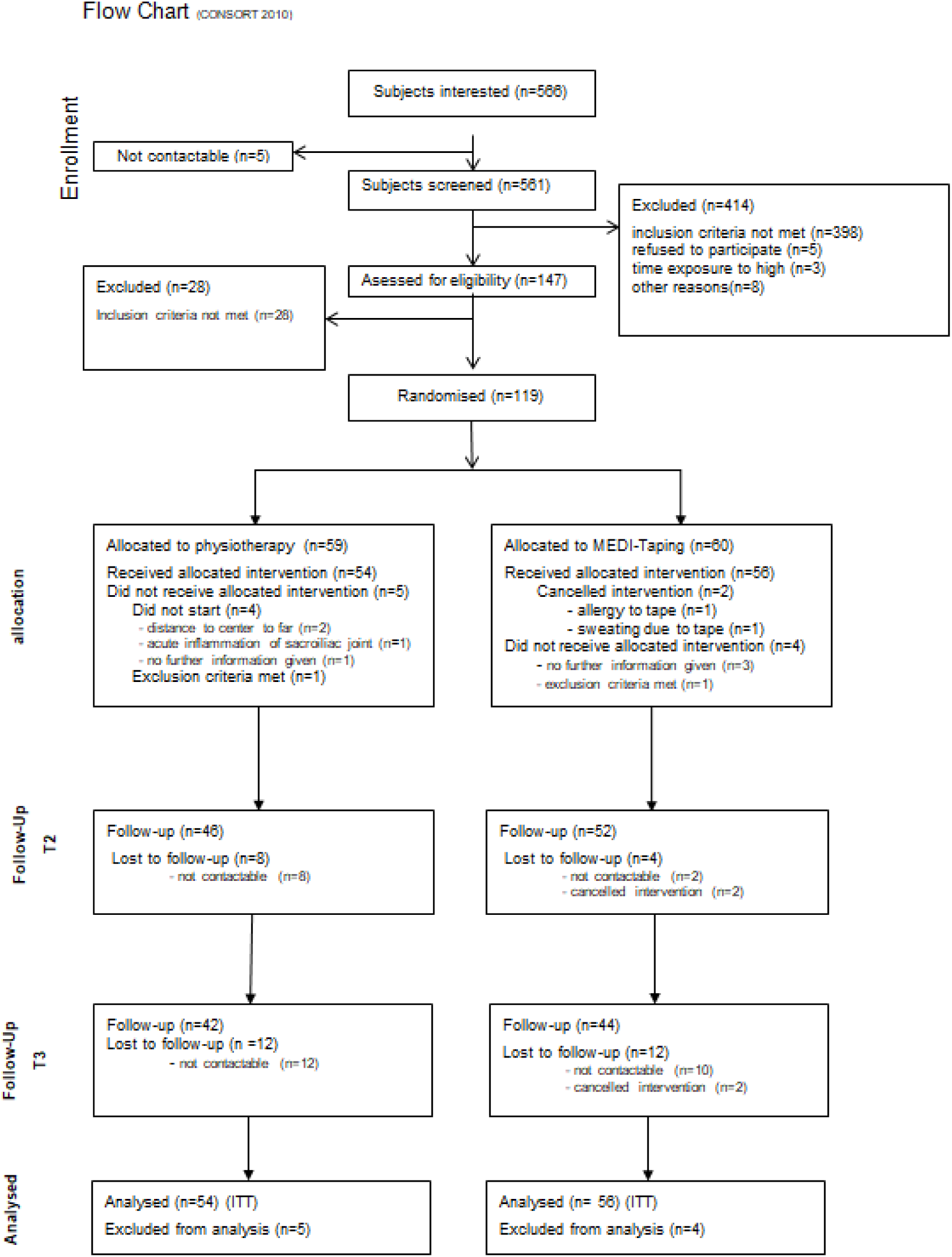
CONSORT patient flow.

Table 1 presents the sociodemographic characteristics of the sample sorted by group as well as the respective p-value of the test for baseline difference, Table 2 the respective clinical baseline characteristics. There were no significant baseline differences for any sociodemographic or clinical variable.

**Table 1.** Sociodemographic characteristics

|  | *Standard Care* |  | *Medi-Taping* |
| --- | --- | --- | --- |
|  | N=54 | N=56 | *p-value* |
| Gender ( % female) | 68.5 | 69.6 |  |
| Age (M;SD) | 52.5 (13.63) | 52.4 (12.81) | .90 |
| Weight | 77.3 (16.80) | 75.4 (18.58) | .96 |
| Height | 171.5 (9.58) | 171.6 (7.93) | .58 |
|  |  |  | .97 |
| Family status % |  |  |  |
| married | 63 | 69.6 | .63 |
| divorced | 14.8 | 8.9 |  |
| single | 11.1 | 16.1 |  |
| living separate | 7.4 | 3.6 |  |
| Living % |  |  |  |
| alone | 22.6 | 23.2 | .49 |
| With partner | 66 | 71.4 |  |
| Shared flat | 7.5 | 5.4 |  |
| With parents | 3.8 | 0 |  |
| Education % |  |  |  |
| In education |  |  | .48 |
| Basic schooling | 22.2 | 14.3 |  |
| GSCE | 37 | 35.7 |  |
| A-level | 40.7 | 50 |  |

**Table 2.**
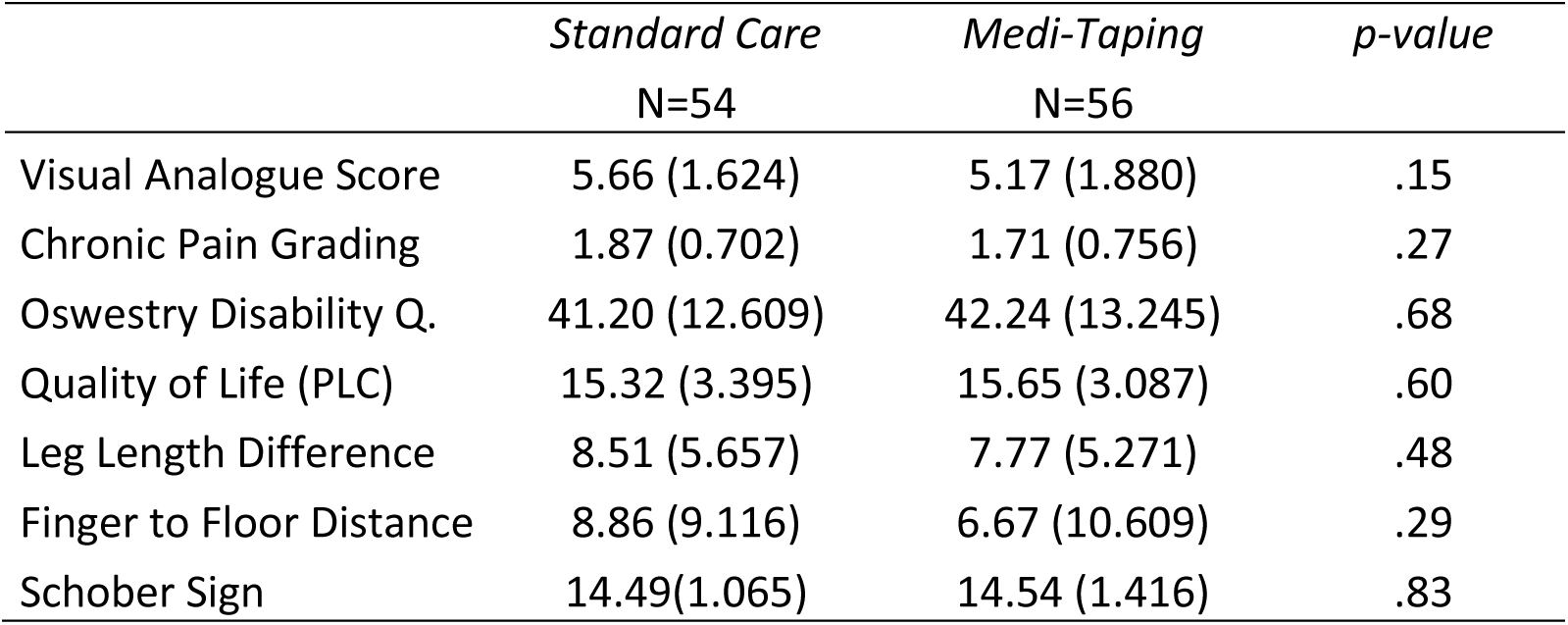
Baseline values of the clinical variables for the two groups; mean values (standard deviations).

|  | *Standard Care*<br>N=54 | *Medi-Taping*<br>N=56 | *p-value* |
| --- | --- | --- | --- |
| Visual Analogue Score | 5.66 (1.624) | 5.17 (1.880) | .15 |
| Chronic Pain Grading | 1.87 (0.702) | 1.71 (0.756) | .27 |
| Oswestry Disability Q. | 41.20 (12.609) | 42.24 (13.245) | .68 |
| Quality of Life (PLC) | 15.32 (3.395) | 15.65 (3.087) | .60 |
| Leg Length Difference | 8.51 (5.657) | 7.77 (5.271) | .48 |
| Finger to Floor Distance | 8.86 (9.116) | 6.67 (10.609) | .29 |
| Schober Sign | 14.49(1.065) | 14.54 (1.416) | .83 |

We also compared the baseline data for the two study centers South (N=59) and North (N=51). Samples were compatible for all variables except pain (VAS) and leg length difference (LLD), with the South sample reporting significant more pain than the North one (South M=5.79, SD=1.4, North M=4.97, SD=2.0, T=2.494, df=108, p=.014). Leg length difference was smaller in the South center (M=5.36, SD=3.15) than in the North center (M=11.34, SD=5.8). This difference was highly significant (T=6.847, df=108, p<.001).

### Hypotheses

All analyses in the results section are based on the ITT-sample with N=110 (Medi-Taping n=56, standard care n=54). Table 3 reports the descriptive data for all variables and all measurements by group.

**Table 3.** Descriptive data for all clinical parameters for the two groups and the three measurement points t1=baseline, t2=end of therapy, t3=follow up (three months) * primary outcomes at t2

|  |  | *t1* |  | *t2* |  | *t3* |  |
| --- | --- | --- | --- | --- | --- | --- | --- |
|  |  | *M* | (*SD*) | *M* | (*SD*) | *M* | (*SD*) |
| VAS – visual analogue Scale* | Standard Care | 5.66 | (1.62) | 4.19 | (2.07) | 4.60 | (2.24) |
|  | Medi-Taping | 5.17 | (1.88) | 3.31 | (2.32) | 3.30 | (2.17) |
| CPGS – Korff pain grading scale* | Standard Care | 1.87 | (0.70) | 1.57 | (0.79) | 1.56 | (0.79) |
|  | Medi-Taping | 1.71 | (0.76) | 1.61 | (0.78) | 1.36 | (0.59) |
| ODQ - Oswestry Disability quest.* | Standard Care | 41.20 | (12.61) | 38.02 | (14.26) | 38.35 | (13.47) |
|  | Medi-Taping | 42.23 | (13.24) | 36.70 | (14.42) | 35.12 | (12.47) |
| PLC – Quality of Life Profile | Standard Care | 15.32 | (3.39) | 15.71 | (3.32) | 15.65 | (3.50) |
|  | Medi-Taping | 15.65 | (3.09) | 17.16 | (3.16) | 16.97 | (3.21) |
| LLD - Leg Length Difference (mm) | Standard Care | 8.51 | (5.66) | 3.39 | (3.17) | 3.37 | (3.36) |
|  | Medi-Taping | 7.77 | (5.27) | 3.14 | (3.89) | 3.42 | (3.15) |
| FTF - Floor to Finger Distance (cm) | Standard Care | 8.86 | (9.12) | 8.62 | (8.88) | 8.86 | (8.74) |
|  | Medi-Taping | 6.67 | (10.61) | 5.79 | (10.15) | 6.08 | (9.60) |
| SS - Schober sign | Standard Care | 14.49 | (1.07) | 14.15 | (0.93) | 14.39 | (0.89) |
|  | Medi-Taping | 14.54 | (1.42) | 13.99 | (1.10) | 14.31 | (1.27) |

The results of the linear models testing group differences at the end of therapy (t2) for the three primary outcome variables can be seen in tables 4 and 5. There were no significant group differences at this time point. We also report interaction terms for the study centers which all proved to be not significant.

**Table 4.** Results of linear models for the primary outcome variables pain (VAS), and functional limitations (ODQ) at t2, baseline values entered as covariates.

| *Variable* | *factor* | *F* | *df* | *p-value* | *part. $\eta^2$* |
| --- | --- | --- | --- | --- | --- |
| VAS | group | 2.718 | 1 | .10 | .025 |
|  | group x center | 1.229 | 1 | .27 | .012 |
| ODQ | group | 2.12 | 1 | .14 | .02 |
|  | group x center | 0.02 | 1 | .88 | .0002 |

**Table 5.** Results of Poisson regression for the primary outcome variable chronic pain grade severity according to CPGS (Korff)

| *Variable* | *Effect* | *Wald Chi2* | *df* | *p-value* | *Estimate* | *Conf Int low* | *Conf Int high* | *Rate Ratio* |
| --- | --- | --- | --- | --- | --- | --- | --- | --- |
| CPGS | group | 0.20 | 1 | .64 | -0.03 | -0.18 | 0.11 | 0.97 |
|  | group*center | 0.04 | 1 | .83 | 0.015 | -0.13 | 0.16 | 1.01 |

Tables 6 and 7 present the results of the same analysis for the secondary outcomes. Here we find a significant difference for health related quality of life (PLC) and for Finger-to-Floor Distance. Patients in the Medi-Taping group improved significantly more in quality of life than patients in the standard care group independent of study center. In Finger-to-Floor Distance patients in the Medi-Taping group had significantly better results, improving by 28% compared with control-group patients; here a center effect was obvious (RR = 1.12).

**Table 6.** Results of RM-ANCOVAs for the secondary outcome variables at t2, baseline values functioned as covariates. PLC = Quality of Life Profile

| *Variable* | *factor* | *F* | *df* | *p-value* | *part. $\eta^2$* |
| --- | --- | --- | --- | --- | --- |
| PLC | group | 8.564 | 1 | .004** | .075 |
|  | group x center | 0.146 | 1 | .70 | .001 |

\*\* p<.01

**Table 7.** Results of Linear Models (Poisson regression) for the secondary outcome variables with respective non-normal distribution

| *Variable* | *Effect* | *Wald Chi<sup>2</sup>* | *df* | *p-value* | *Estimate* | *Conf Int low* | *Conf Int high* | *Rate Ratio* |
| --- | --- | --- | --- | --- | --- | --- | --- | --- |
| LLD | group | 0.75 | 1 | .38 | 0.047 | -0.059 | 0.154 | 1.048 |
|  | group*center | 6.29 | 1 | .012 | 0.136 | 0.03 | 0.24 | 1.145 |
| Schober | group | 0.09 | 1 | .76 | 0.007 | -0.04 | 0.05 | 1.007 |
|  | group*center | 0.12 | 1 | .72 | -0.009 | -0.06 | 0.04 | 0.991 |
| FTF | group | 45.97 | 1 | <.00001 | 0.258 | 0.18 | 0.33 | 1.29 |
|  | Group*center | 10.02 | 1 | .0015 | 0.115 | 0.044 | 0.187 | 1.12 |

LLD: leg length difference; FTF: Finger to floor distance

For the assessment of the follow-up data we computed a repeated measurement ANCOVA over the post-treatment and follow-up measurement points with baseline as covariate. The results for both primary and secondary outcomes can be seen in Table 8. We report the relevant interaction terms for time x group. With VAS-Pain rating there was a significant interaction between time and center (F = 3.1, p^corrected^ = .048) indicating that patients in one center did better over time than in the other. No other interactions between center and time were detectable. Figures 2 to 4 illustrate the time course of the scores for VAS, Oswestry Disability Score and Quality of Life.

**Table 8.** Results of Repeated Measurement ANOVA: Interaction Terms for Group*Time Interaction with Greenhouse-Geisser Corrected Degrees of Freedom and according p-Values

|  | *F* | *korr. Df* | *p* | *part. $\eta^2$* |
| --- | --- | --- | --- | --- |
| Visual Analogue Scale | 1.656 | 1.972 | .19 | .015 |
| Oswestry Disability Questionnaire | 2.9 | 1.90 | .060 | .026 |
| Quality of Life (PLC) | 2.998 | 1.841 | .057 | .028 |

**Figure 2-4.**
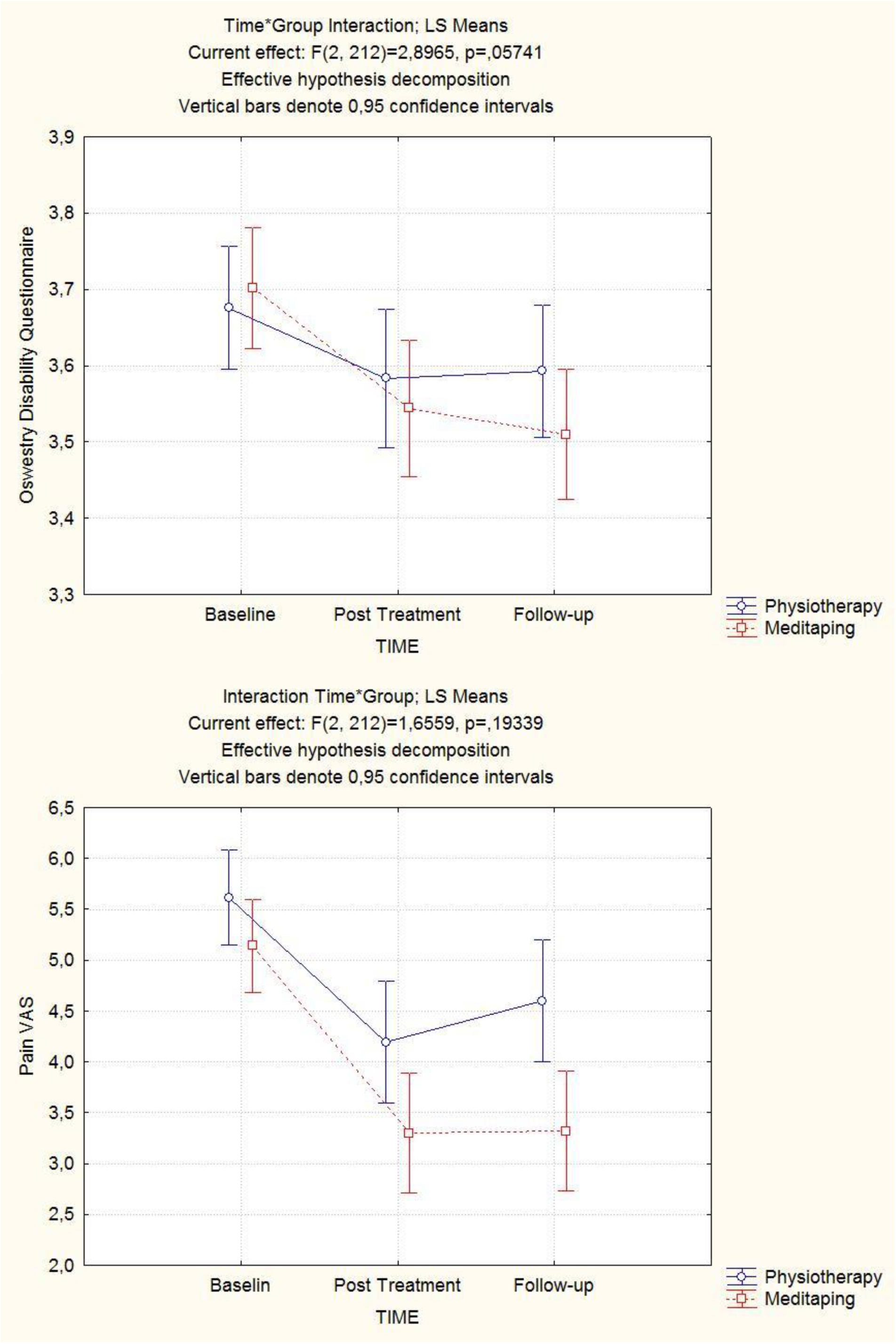
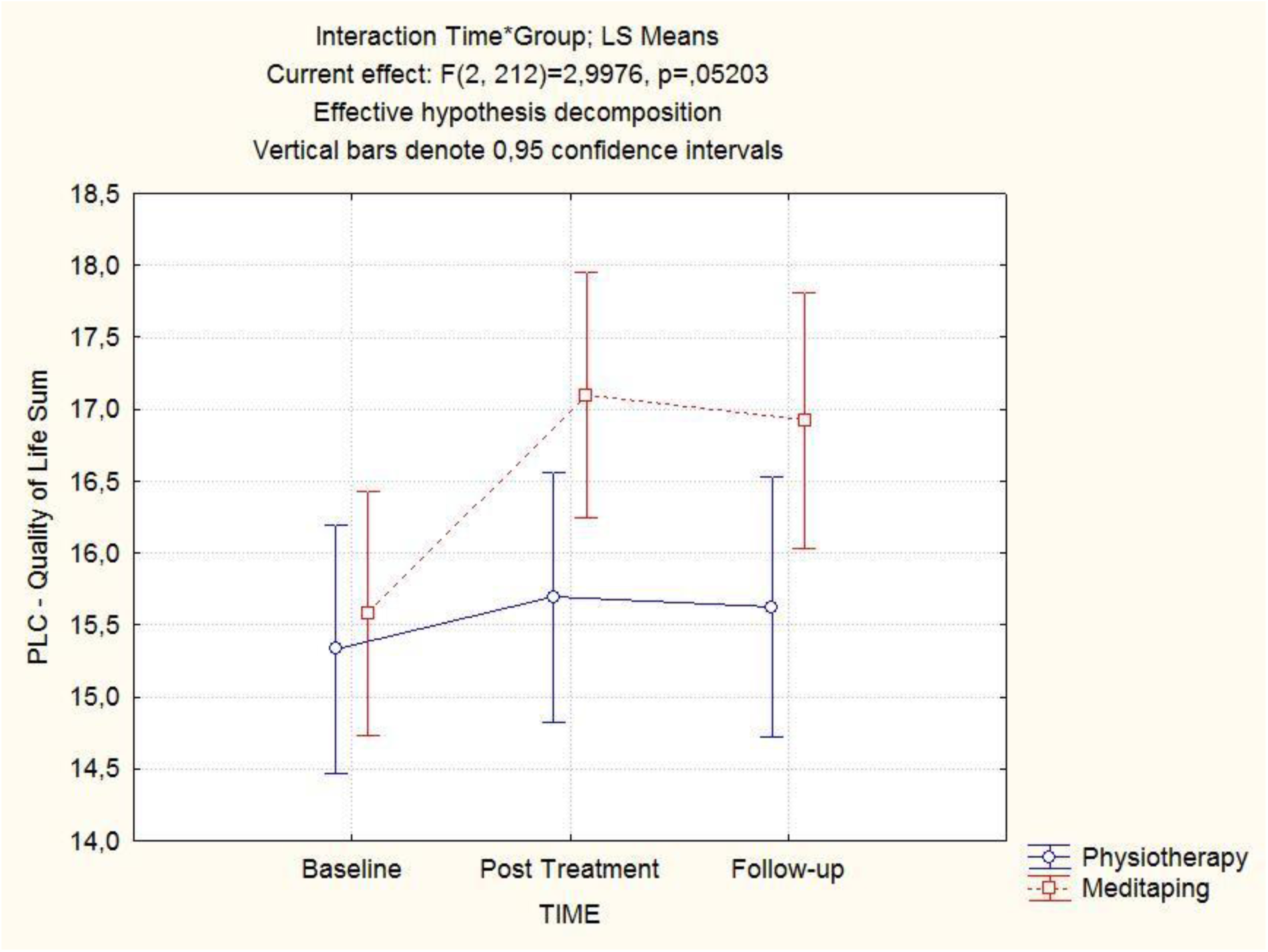
Interaction Plots from RM-Anovas to Illustrate the Time Course of Oswestry Disability Score (fig. 2), VAS (fig.3), and Quality of Life (fig. 4)

For the Poisson-distributed variables we computed Poisson-regression models for the follow-up score as dependent variable with the baseline and post-treatment score as covariates and group and center as categorical predictors (Table 9).

**Table 9.** Results of Linear Models (Poisson regression) for the secondary outcome variables; dependent variable score at follow-up; predictors scores at baseline and post-treatment, group and center

| Variable | Effect | Wald<br>Chi<sup>2</sup> | df | p-value | Estimate | Conf<br>Int low | Conf<br>Int high | Rate<br>Ratio |
| --- | --- | --- | --- | --- | --- | --- | --- | --- |
| LLD | group | 0.17 | 1 | .68 | -0.02 | -0.12 | 0.08 | 0.98 |
| Schober | group | 0.006 | 1 | .93 | 0.007 | -0.051 | 0.048 | 0.998 |
| FTF | group | 36,31 | 1 | <.00001 | 0.222 | 0.15 | 0.29 | 1.25 |
| Korff | Group | 0.58 | 1 | .44 | 0.06 | -0.09 | 0.22 | 1.06 |

LLD: Leg length difference; FTF: Finger to floor distance

The interaction plots (Figures 2-4) illustrate that in each case the Medi-Taping group was better than the physiotherapy group and the improvements were more sustainable in the Medi-Taping group. We therefore conducted a multivariate analysis of these three variables together over both post-treatment and follow-up with baseline as a predictor as a kind of post-hoc exploratory analysis. Here, a clearly significant time*group interaction can be seen (Wilk’s lamba = 0.88; p = .004; partial eta^2^ = .12). This illustrates that our a-priori effect size assumption was too optimistic and the study suffered from a power problem.

### Within-group effect sizes

In order to also document within-group changes as a measure of overall clinical improvement we computed pre-post effect sizes from baseline to the two endpoints t2 and t3 (Table 10).

**Table 10.** pre-post effect sizes (Cohen’s d) for changes from t1 to t2 and from t1 to t3 for both study arms.

| *Variable* | *t1-t2* |  | *t1-t3* |  |
| --- | --- | --- | --- | --- |
|  | *Medi-Taping* | *Standard care* | *Medi-Taping* | *Standard care* |
| Visual Analogue Scale | 0.89 | 0.80 | 0.92 | 0.55 |
| CPGS (Korff) | 0.13 | 0.40 | 0.52 | 0.42 |
| Oswestry Disability Questionnaire (ODQ) | 0.40 | 0.24 | 0.55 | 0.22 |
| Quality of Life (PLC) | 0.48 | 0.12 | 0.42 | 0.10 |
| Leg Length Difference (LLD) | 1.01 | 1.16 | 1.03 | 1.14 |
| Finger to Floor Distance (FFT) | 0.08 | 0.03 | 0.06 | 0.00 |
| Schober Sign | 0.44 | 0.34 | 0.17 | 0.10 |

Within-group effect sizes were large for pain reduction, for the Medi-Taping group at both time points and for the physiotherapy group at post-treatment. Effect sizes for functionality and the Korff-pain grading were medium sized with Medi-Taping at follow-up and small with physiotherapy. Both treatments achieved large effect sizes for leg-length difference at both measurement points. Between group effect sizes for VAS pain rating (difference-score between baseline and follow-up) was d = 0.34 and thus small.

## Discussion

This pragmatic, examiner blind randomized controlled trial of Medi-Taping versus physiotherapy in patients with chronic low back pain was the first of its kind. It was powered to detect medium-sized between group effects in favor of Medi-Taping. Our primary analysis failed to confirm this difference, as none of the primary outcomes showed a clearly significant effect in favor of the experimental treatment. Although all effects were pointing in the anticipated direction with patients in the Medi-Taping group doing better, the significance levels of the pain and the disability variables showed only tendencies. Some of the secondary outcome variables showed an effect: Quality of life improved significantly more in the Medi-Taping group with a small to medium sized effect of about 7% variance explained. And the finger-to-floor distance that measures functionality improved clearly and significantly more in the Medi-Taping group with a rate ratio of 1.29 indicating that a patient in the Medi-Taping group had on average a 29% better chance of improvement. This result was confirmed by the secondary analysis, a repeated measure analysis of variance of all variables, where the Oswestry Disability Score and the Quality of Life Score (PLC) just missed significance, and the Finger-to-Floor Distance also was highly significant. The trend illustrated by the interaction plots (Figures 2-4) and the multivariate analysis confirm that the Medi-Taping group is better in tendency, but the effect was much too small to be picked up by our study, and thus the study suffered from a power problem.

Patients in both groups profit from the treatments, as is demonstrated by significant time effects of RM-ANOVAs and the within-group effect sizes. Thus, the strong control of the physiotherapy treatment was difficult to outrun with a trial of this size. An effect size of the magnitude found in our study (d = 0.37 for VAS at follow-up) would require 155 patients in each group to meet a power goal of 90% statistical power in a superiority trial and would thus have to be three times as large as this trial.

One should, however, consider that the physiotherapy treatment offered in this trial was more intensive and thus more time-consuming and cost intensive than the Medi-Taping treatment, which was shorter and needed fewer sessions.

The trial was pragmatic. Although all results provided by clinicians are blind, patients could not be blinded, and the taping-treatment would not lend itself easily to any form of blinding control. Hence one might suspect reporting bias in the patient outcomes. However, as patients knew that they would be receiving one of two potentially effective treatments and clinicians were blinded, we reckon that reporting bias was not a problem, especially since the difference between groups in self-reported measures was not larger than in clinician reported outcomes. The patients in our study were certainly comparable to patients in general practice. They had a chronic problem. Their pain was partially disabling them in their function and it was of medium severity. Hence, our trial can be generalized to a community sample, and both treatments can be useful for chronic pain.

The most sensitive outcome proved to be finger-to-floor distance. This is a robust and objective measure of functionality. For a follow-up study, this outcome should be kept, and the two other clinician rated outcomes might be dropped. The patient reported outcomes we used are standard ones. Only the quality of life scale really differentiated, while the Oswestry disability scale produced only small effects. This might have to do with the fact that the patients in our sample were not really severely compromised. Perhaps some other instrument, such as the Orebrö scale (Linton and Halldén 1998; Schmidt et al., 2014) might be more useful and sensitive.

The intervention studied here is certainly apar with other effective chronic pain treatments. The physiotherapy offered here was a strong multimodal package with elements of mobilization and patient education. This is testified by the large pre-post effect sizes.

One of the studies with the largest effects was a trial of Yoga against standard care which found a significant improvement in the Roland Morris Disability Score, a score comparable to the Oswestry score used here, of 2 points for the Yoga group and 0,48 points for the standard care group. Although this measure was documented after a year it seems that the improvements found by our study after 3 months of 3 points for the physiotherapy group and 7 points for the Medi-Taping group compare well against this study. Only a comparatively complex and long term treatment, Alexander technique, seems to be more efficient (Little et al., 2008).

### Our study had limitations

It was the first of its kind and hence all we could go by were educated guesses and clinical impressions which are notoriously unreliable, as documented once more by our too optimistic power calculation. While we operated with two centers and thus had the benefit of some limited generalisability, ideally more centers should have been included. Perhaps a more realistic design would have been a non-inferiority trial, which, however, would require a considerably larger sample size (Committee for Proprietary Medicinal Products (CPMP) 2000; Wiens 2002).

## Conclusions

We conclude from our pragmatic, partially blinded randomized study that Medi-Taping, a purported way of correcting pelvic obliquity and chronic tension, is a treatment modality similar in effectiveness in treating chronic low back pain as a complex physiotherapy and patient education program. There are indications that Medi-Taping is better long term and improves quality of life more than standard physiotherapy, but these indications are tentative and require replication in an adequately powered study which would need approximately 300 patients to confirm our findings.

### Clinical Study Registration

This trial was registered under the Number DRKS00017051 in the German Register of Clinical Trials (Deutsches Register Klinischer Studien).

## Data Availability

data can be obtained from the corresponding author.

## References

Ärztliches Zentrum für Qualität in der Medizin (ÄZQ).Chronischer Kreuzschmerz. Aktiv gegen chronischen Kreuzschmerz: Patientenleitlinie zur Nationalen Versorgungsleitlinie Kreuzschmerz (Chronic low back pain: Active against back pain: Patient guidelines for the National Guideline Back Pain). Berlin: Bundesärztekammer; 2011.

Ayanniyi O, Raji FS, Adegoke BOA. Prevalence of asymptomatic sacroiliac joint dysfunction and its association with leg length discrepancies in male students in selected junior secondary schools in IbadanAfrican Journal of Medicine and Medical Sciences 2008;37.

Bordoni B and Marelli F. Emotions in motion: myofascial interoception. Complementary Medicine Research 2017;24: 110–113.

Chou R, Deyo R, Friedly J, Skelly A, Hashimoto R, Weimer M, Fu R, Dana T, Kraegel P, Griffin J, Grusing S, Brodt ED. Nonpharmacologic therapies for low back pain: A systematic review for an american college of physicians clinical practice guideline. Annals of Internal Medicine 2017;166: 493–505.

Committee for Proprietary Medicinal Products (CPMP).Points to consider on switching between superiority and non-inferiority In: E-EAftEoM Products, editor London: EMA; 2000.

Coxib and traditional NSAID Trialists’ (CNT) Collaboration. : Vascular and upper gastrointestinal effects of non-steroidal anti-inflammatory drugs: meta-analyses of individual participant data from randomised trials. The Lancet 2013;382: 769–779.

Critchley DJ, Ratcliffe J, Noonan S, Jones RH, Hurley MV. : Effectiveness and cost-effectiveness of three types of physiotherapy used to reduce chronic low back pain disability: A pragmatic randomized trial with economic evaluation. Spine 2007;32: 1474–1481.

Dowell D, Haegerich TM, Chou R. CDC guideline for prescribing opioids for chronic pain - United States 2016. Journal of the American Medical Association 2016;online first.

Dworkin RH, Turk DC, Farrar JT, Haythornthwaite JA, Jensen MP, Katz NP, Kerns RD, Stucki G, Allen RR, Bellamy N, Carr DB, Chandler J, Cowan P, Dionne R, Galer BS, Hertz S, Jadad AR, Kramer LD, Manning DC, Martin S, McCormick CG, McDermott MP, McGrath P, Quessey S, Rappaport BA, Robbins W, Robinson JP, Rothman M, Royal MA, Simon L, Stauffer JW, Stein W, Tollett J, Wernicke J, Witter J. Core outcome measures for chronic pain clinical trials: IMMPACT recommentatioins. Pain 2005;113: 9–19.

Ekman M, Jönhagen S, Hunsche E, Jönsson L. Burden of illness of chronic low back pain in Sweden: A cross-sectional, retrospective study in primary care setting. Spine 2005;30: 1777–1785.

Fairbank J, Davies J, Couper J, al. e. The Oswestry low back pain disability questionnare. Physiotherapy 1980;66: 271–273.

Farioli A, Mattioli S, Quaglieri A, Curti S, Violante FS, Coggon D. Musculoskeletal pain in Europe: role of personal, occupational and social risk factors. Scandinavian Journal of Work, Environment & Health 2014;40: 36–46.

Gaul C, Mette E, Schmidt T, Grond S. Praxistauglichkeit einer deutschen Version des “Oswestry Low Back Pain Disability Questionnaire”. Ein Fragebogen zur Beeinträchtigung durch Rückenschmerzen. Der Schmerz 2008;22 51–58.

Global Burden of Disease Study 2013 Collaborators. Global, regional, and national incidence, prevalence, and years lived with disability for 301 acute and chronic diseases and injuries in 188 countries, 1990-2013;2013: a systematic analysis for the Global Burden of Disease Study 2013. The Lancet 2015;386: 743–800.

Gor AP and Saksena M. Adverse drug reactions of nonsteroidal anti-inflammatory drugs in orthopedic patients. Journal of Pharmacology and Pharmacotherapeutics 2011;2: 26–29.

Holm S. A simple sequentially rejective multiple test procedure. Scandinavian Journal of Statistics 1979;6: 65–70.

Huskisson E. Measurement of pain. Lancet 1974;4: 1127–1131.

Huskisson E. Visual analogue scales. In; 1983.

Ilic B, Nikolic A, Ilic D. Efficiency of kinesio taping in prevention and rehabilitation of sport injuries. Sportlogia 2017;13: 53-65.

Ingber DE, Wang N, Stamenović D. Tensegrity, cellular biophysics, and the mechanics of living systems. Reports on Progress in Physics 2014;77: 046603.

Kase K, Hashimoto T, Tomoki O. Development of kinesio taping perfect manual. Kinesio Taping Association 1996;6: 117–118.

Klasen BW, Hallner D, Schaub C, Willburger R, Hasenbring M. Validation and reliability of the German version of the Chronic Pain Grade questionnaire in primary care back pain patients. GMS Psycho-Social-Medicine 2004;1: 1–14.

Kuijpers T, van Middelkoop M, Rubinstein SM, Ostelo R, Verhagen A, Koes BW, van Tulder MW. A systematic review on the effectiveness of pharmacological interventions for chronic non-specific low-back pain. European Spine Journal 2011;20: 40–50.

Langevin HM, Stevens-Tuttle D, Fox JR, Badger GJ, Bouffard NA, Krag MH, Wu J, Henry SM. Ultrasound evidence of altered lumbar connective tissue structure in human subjects with chronic low back pain. BMC Musculoskeletal Disorders 2009;10: 151.

Lim ECW and Tay MGX. Kinesio taping in musculoskeletal pain and disability that lasts for more than 4 weeks: is it time to peel off the tape and throw it out with the sweat? A systematic review with meta-analysis focused on pain and also method of tape application. British Journal of Sports Medicine 2015;49: 1558–1566.

Linton SJ and Halldén K. Can we screen for problematic back pain? A screening questionnaire for acute and subacute back pain. Clinical Journal of Pain 1998;14: 209–215.

Little P, Lewith G, Webley F, Evans MR, Beattie A, Middleton K, Barnett J, Ballard K, Oxford F, Smith PK, Yardley L, Hollinghurst S, Sharp D. Randomised controlled trial of Alexander technuique lessons, exercise and massage (ATEAM) for chronic recurrent back pain. British Medical Journal 2008;337: a884.

Moll JM and Wright V. Normal range of spinal mobility. An objective clinical study. Annals of the Rheumatic Diseases 1971;30: 381–386.

Nelson NL. Kinesio taping for chronic low back pain: A systematic review. Journal of Bodywork and Movement Therapies 2016;20: 672–681.

Nijs J, Clark J, Malfliet A, Ickmans K, Voogt L, Don S, den Bandt H, Goubert D, Kregel J, Coppieters I, Dankaerts W. In the spine or in the brain? Recent advances in pain neuroscience applied in the intervention for low back pain. Clinical and Experimental Rheumatology 2017;35: S108–S115.

Ofner M, Kastner A, Schwarzl G, Schwameder H, Alexander N, Strutzenberger G, Walach H. RegentK and physiotherapy support knee function after anterior cruciate ligament rupture without surgery after 1yYear: A randomized controlled trial. Complementary Medicine Research 2017;online first.

Parreira PdCS, Costa LdCM, Hespanhol Junior LC, Lopes AD, Costa LOP. Current evidence does not support the use of Kinesio Taping in clinical practice: a systematic review. Journal of Physiotherapy 2014;60: 31–39.

Qaseem A, Wilt TJ, McLean RM, Forciea M, Physicians; ftCGCotACo. Noninvasive treatments for acute, subacute, and chronic low back pain: A clinical practice guideline from the american college of physicians. Annals of Internal Medicine 2017;166: 514–530.

Raspe H-H.Rückenschmerzen [Back pain]. Gesundheitsberichterstattung des BundesBerlin: Robert-Koch-Institut; 2012.

Saper RB, Lemaster C, Delitto A, Sherman KJ, Herman PM, Sadikova E, Stevans J, Keosaian JE, Cerrada CJ, Femia AE, Roseen EJ, Gardiner P, Barnett KG, Faulkner C, Weinberg J. Yoga, physical therapy, or education for chronic low back pain: A randomized noninferiority trial. Annals of Internal Medicine 2017;167: 85–94.

Schleip R, Findley TW, Chaitow L, Huijing PA. Fascia: The Tensional Network of the Human Body.London: Churchill Livingstone/Elsevier; 2012.

Schmidt C, Raspe H, Pfingsten M. Back pain in the German adult population. Prevalence, severity, and sociodemographic correlates in a multiregional survey. Spine 2007;32: 2005–2011.

Schmidt CO, Lindena G, Pfingsten M, Kohlmann T, Chenot J-F. Vergleich zweier Screening-Fragebogen für Patienten mit Rückenschmerzen. Der Schmerz 2014;28: 365–373.

Schober P. Lendenwirbelsäule und Kreuzschmerzen (The lumbar vertebral column in backache). Münchener Medizinische Wochenschrift 1937;84: 336–338.

Siegrist J, Broer M, Junge A. Profil der Lebensqualität bei chronisch Kranken (PLC). Göttingen: Hogrefe. 1994.

Spijker-Huiges A, Groenhof F, Winters JC, van Wijhe M, Groenier KH, van der Meer K. Radiating low back pain in general practice: Incidence, prevalence, diagnosis, and long-term clinical course of illness. Scandinavian Journal of Primary Health Care 2015;33: 27–32.

Tilbrook HE, Cox H, Hewitt CE, Kang’ombe AR, Chuang L-H, Jayakody S, Aplin JD, Semlyen A, Trewhela A, Watt I, Torgerson DJ. Yoga for chronic low back pain. Annals of Internal Medicine 2011;155: 569–578.

Tozzi P. A unifying neuro-fasciagenic model of somatic dysfunction - underlying mechanisms and treatment - Part 2. Journal of Bodywork and Movement Therapies 2015a;19: 526–543.

Tozzi P. A unifying neuro-fasciagenic model of somatic dysfunction - underlying mechanisms and treatment - part I.. Journal of Bodywork and Movement Therapies 2015b;19: 310–326.

van Middelkoop M, Rubinstein SM, Kuijpers T, Verhagen AP, Ostelo R, Koes BW, van Tulder MW. A systematic review on the effectiveness of physical and rehabilitation interventions for chronic non-specific low back pain. European Spine Journal 2011;20: 19–39.

Vanti C, Bertozzi L, Gardenghi I, Turoni F, Guccione AA, Pillastrini P. Effect of Taping on Spinal Pain and Disability: Systematic Review and Meta-Analysis of Randomized Trials. Physical Therapy 2015;95: 493–506.

Von Korff M, Ormel J, Keefe FJ, Dworkin SF. Grading the severity of chronic pain. Pain 1992;50: 133–149.

Wellington J. Noninvasive and alternative management of chronic low back pain (efficacy and outcomes). Neuromodulation: Technology at the Neural Interface 2014;17: 24–30.

Wiens BL. Choosing an equivalence limit for noninferiority or equivalence studies. Controlled Clinical Trials 2002;23: 2–14.

Williams S, Whatman C, Hume PA, Sheerin K. Kinesio taping in treatment and prevention of sports injuries: A meta-analysis of the evidence for its effectiveness. Sports Medicine 2012 42: 153–164.

